# Modeling the influence of vaccine administration on COVID-19 testing strategies

**DOI:** 10.1101/2021.10.14.21265035

**Authors:** Jonathan E. Forde, Stanca M. Ciupe

## Abstract

Vaccination is considered the best strategy for limiting and eliminating the COVID-19 pandemic. The success of this strategy relies on the rate of vaccine deployment and acceptance across the globe. As these efforts are being conducted, the severe acute respiratory syndrome coronavirus 2 (SARS-CoV-2) virus is continuously mutating, which leads to the emergence of variants with increased transmissibility, virulence, and lower response the vaccines. One important question is whether surveillance testing is still needed in order to limit SARS-CoV-2 transmission in an increasingly vaccinated population. In this study, we developed a multi-scale mathematical model of SARS-CoV-2 transmission in a vaccinated population and used it to predict the role of testing in an outbreak with alpha and delta variants. We found that, when the alpha variant is dominant, testing is effective when vaccination levels are low to moderate and its impact is diminished when vaccination levels are high. When the delta variant is dominant, widespread vaccination is necessary in order to prevent significant outbreaks. When only moderate vaccination can be achieved, frequent testing can significantly reduce the cumulative size of delta variant outbreak, with the impact of testing having maximum effects when focused on the non-vaccinated population.

## 1 Introduction

The emergence in 2019 of the novel coronavirus-2 severe acute respiratory syndrome (SARS-CoV-2) in Wuhan, China [42] has had devastating global consequences including loss of lives [6], strained healthcare systems [20, 26, 38], and economic recessions. Protective measures, such as masking, physical distancing, testing, contact tracing, quarantine, and isolation, while effective when applied rigorously, have proved insufficient in limiting the spread of SARS-CoV-2 [23, 30, 35]. Development of COVID-19 vaccines has been the main focus of public health organizations and pharmaceutical companies across the world [10, 25, 36].

As of August 29, 2021 twenty-two vaccines have been approved for emergency or full use by at least one WHO-recognized regulatory authority [7]. The U.S. Food and Drug Administration (FDA) has approved two mRNA (Pfizer-BioNTech and Moderna) and one viral vector-based (Janssen) COVID-19 vaccines [5], which have consistently demonstrated effectiveness against disease and increased protection against infection [9, 18, 39, 40]. Despite early control in highly vaccinated communities, vaccine shortage in low-access countries and vaccine hesitancy in high-access countries or regions, has led to selection of new variants [1], which might overcome vaccine-induced immunity [17, 28].

Delta variant (B.1.617.2), a highly contagious SARS-CoV-2 virus strain, was first identified in India in December 2020 [15], and has since been labeled a variant of concern [11]. It was first identified in the United States in March 2021 and accounted for 98.4% of new infections by September 23, 2021 [1]. The Centers for Disease Control and Prevention (CDC) described delta to be as contagious as chickenpox in an internal document (according to The New York Times). That means that the transmissibility rate is 1.5-2 times higher than that of the alpha variant (B.1.1.7). Recent studies, measuring the effectiveness of vaccines against the transmission of the delta variant, found a reduction in the Pfizer-BioNTech effectiveness to 33.5% and 88% after one and two doses, compared to 49% and 94% effectiveness against the transmission of the alpha variant [28]. CDC defines vaccine breakthrough infection as the detection of SARS-CoV-2 RNA or antigen in a respiratory specimen collected from a person ≥ 14 days after receipt of all recommended doses of an FDA-authorized COVID-19 vaccine [12]. The increased transmissibility of the delta variant and the observed waning in vaccine effectiveness against infection highlight the critical importance of vaccinating an entire community and of offering boosters to individuals 65 years and older vaccinated six months ago [3]. With vaccines not yet approved for children aged 11 or younger, it is also important to adhere to rigorous COVID-19 prevention strategies such as masking and testing. One question of importance to the public health authorities, and the goal of this study, is to determine the best strategy for using testing in an increasingly vaccinated population in order to limit transmission.

SARS CoV-2 diagnostic and surveillance testing is an important intervention tool for controlling transmission. While still widely used in monitoring non-vaccinated population, little is known about the potential benefit of testing vaccinated population. Given limited resources, it is important to identify who to test in order to most effectively control transmission at minimal cost.

In this study, we develop a multi-scale mathematical model to examine the effect of testing on blocking SARS CoV-2 infections in populations vaccinated with the Pfizer-BioNTech vaccine [13, 16, 34]. We expand our previous work, which connected a within-host model for the time of SARS CoV-2 infectiousness onset based on an individual virus dynamics and a between-host model for the transmission at the population level [16]. In particular, we incorporate a variable for the population vaccinated with the Pfizer-BioNTech vaccine and a population of vaccinated individuals that become infected. We connect the vaccine effectiveness in preventing infection with the age of vaccination and the effect of the vaccine on reported individual virus profile when a breakthrough infection occurs [27]. We are interested in determining how vaccine prevalence combined with surveillance testing can help reduce an infectious event with alpha and delta variants.

## 2 Materials and Methods

We model the interaction between a susceptible class *S*(*t*), vaccinated class *v*(*t, α*), infected class of asymptomatic individuals *i*_*a*_(*τ, t*), infected class of symptomatic individuals *i*_*s*_(*τ, t*), infected class of vaccinated individuals *i*_*v*_(*τ, t*), individuals recovered from natural infection *R*(*t*), individuals vaccinated after natural infection *R*_*v*1_(*t*) and individuals recovered after being vaccinated and then infected *R*_*v*2_. The independent variables are the age of infection in an individual *τ*, the age of vaccination in an individual *α*, and the time-since-outbreak in the population *t*. The parameters are the transmission rate *β*, the infection weighting functions *λ*_*j*_, the birth rate *b*, the death rate *µ*, the disease-induced mortality rates *m*_*j*_, the vaccination rate *v*, and the degree of protection after vaccine administration *η*. Moreover, we consider testing in both vaccinated and non-vaccinated populations, with a testing capacity of *C* tests per day, leading to case detection rates *r*_*j*_, with *j* ∈ *{a, s, v}*.

As in our earlier study [16], we assume that an individual’s infection status is given by its virus profile at age of infection *τ*. In particular, given virus profiles for infected individuals, we link test positivity to the ages of infection during which virus load is above the sensitivity threshold. Similarly, we determine the ages of infection during which the virus load is high enough to allow transmission. We define

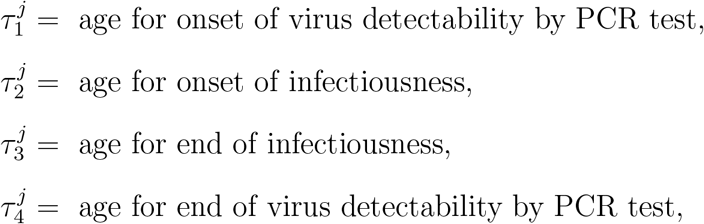

where *j* ∈ *{a, s, v}*. The infectivity weighting functions *λ*_*j*_ are

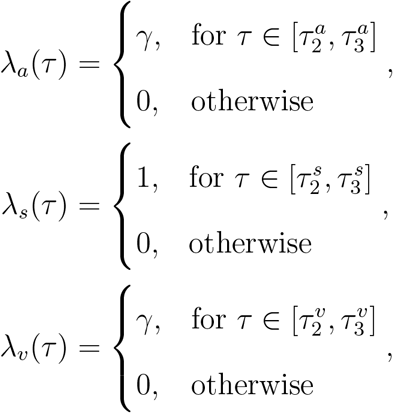

where parameter 0 < *γ* < 1 represents the relative infectiousness of asymptomatic carriers, in comparison with symptomatic carriers. For all infected classes, 0 ≤ *τ* ≤ *τ*_*q*_. For *τ* > *τ*_*q*_ infections are considered resolved, and recovered individual are not susceptible to reinfection. Moreover, we assume that vaccinated individuals who are subsequently infected have asymptomatic disease.

### Daily testing rate

As before [16], we define a daily per capita testing rate, *ρ*, corresponding to an overall testing capacity of *C* tests per day at time *t* in a population of size *N* (*t*) to be given by

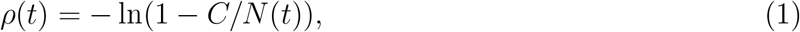

with *C* < *N* (*t*). The case detection rate functions *r*_*j*_(*τ, t*) become

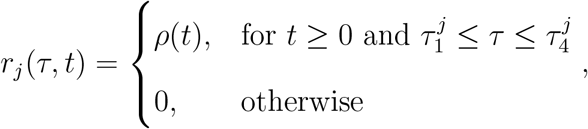

where *j* ∈ *{a, s, v}*. We assume test return delay of *ℓ* days.

If testing is administered to only non-vaccinated individuals, then the relationship between testing capacity *C* and daily per capita testing rate, *ρ* is given by

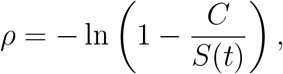

with *C* < *S*(*t*).

If testing is administered to both vaccinated and non-vaccinated individuals, then the relationship between testing capacity *C* and daily per capita testing rate, *ρ* is given by

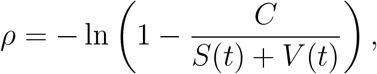

with *C* < *S*(*t*).

### Model equations

On the domain *t* ≥ 0, 0 ≤ *τ* ≤ *τ*_*q*_, and *α* ≥ 0, the between-host model equations under combined vaccination and testing are

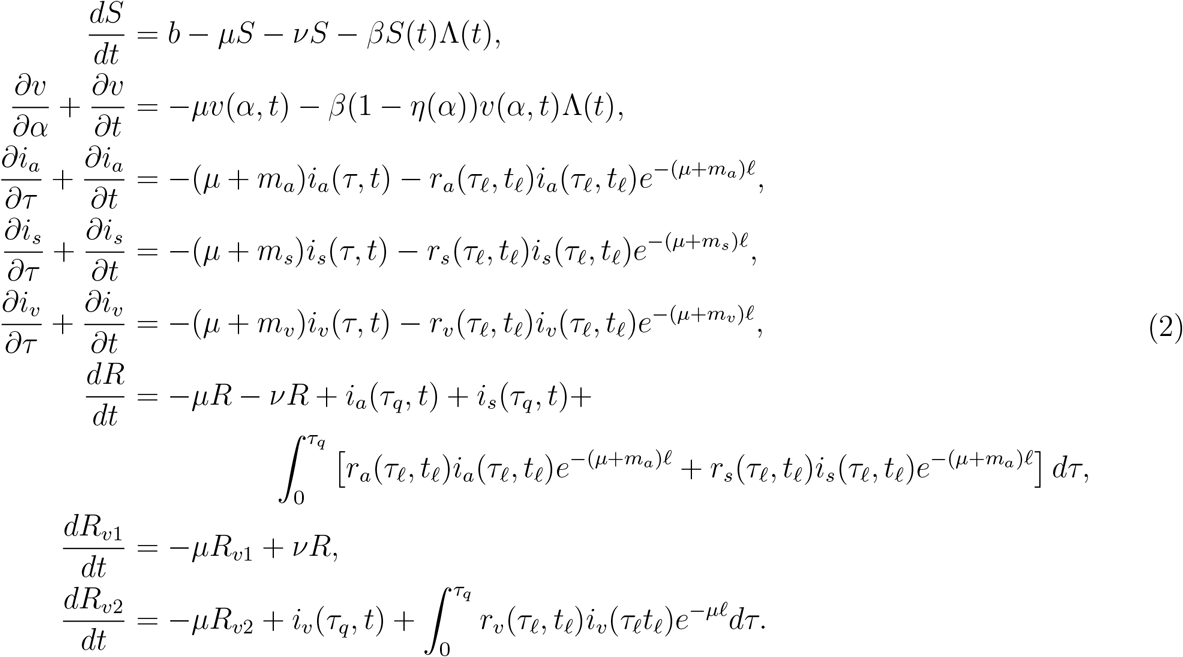

where

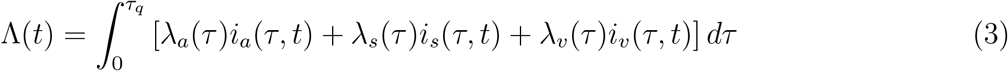

is the weighted infectious population. The subscript *ℓ* represents a delay of *ℓ* days, so *τ*_*ℓ*_ = *τ* − *ℓ, t*_*ℓ*_ = *t* − *ℓ*.

The boundary and initial conditions are

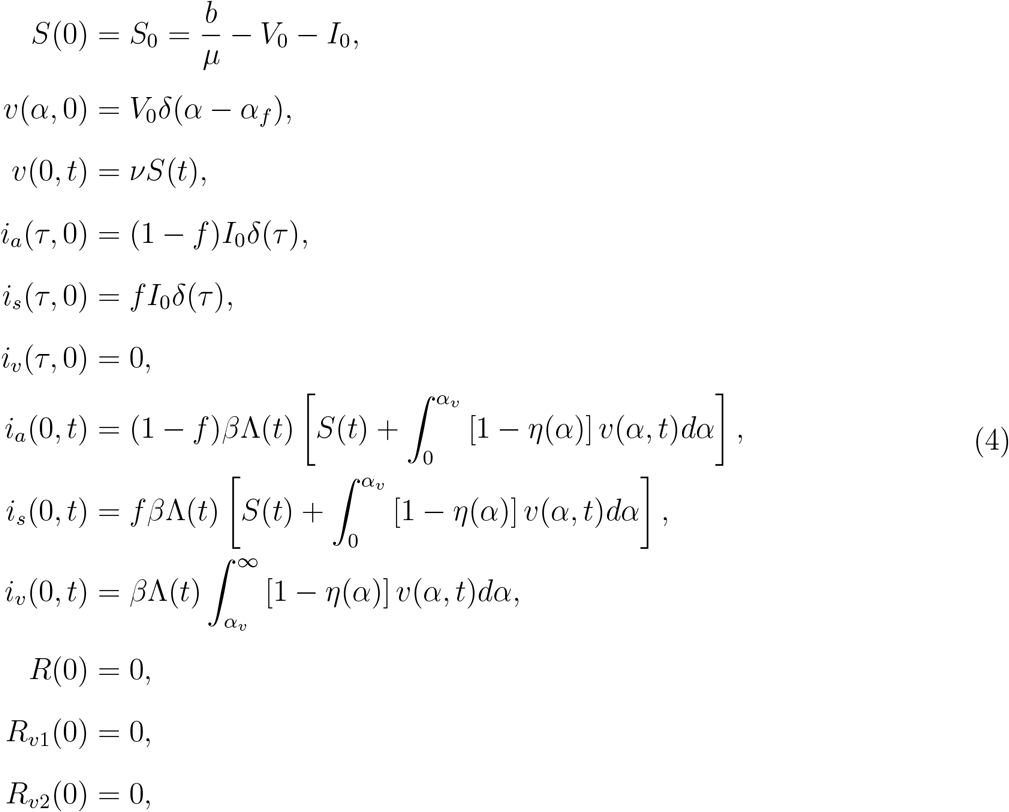

where *f* is the fraction of infections that are symptomatic, *α*_*v*_ is the age of vaccination where an individual’s virus load is reduced if infected, and *α*_*f*_ is the age of vaccination where vaccines provide full protection. For *t* < *α*_*v*_, vaccinated individuals are equivalent to susceptible individuals. Parameters *{β, µ, m*_*a*_, *m*_*s*_, *f, α*_*v*_*}* are taken from literature, and *δ* is the Dirac delta function.

### 2.1 Cumulative Statistics

In order to compare model results with commonly tabulated public health data, we define several cumulative population statistics derived from the model state variables.

The cumulative number of cases up to time *t*, Σ(*t*), is given by the equation

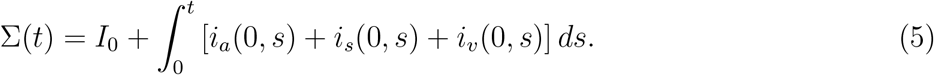

The cumulative number of positive case detections at time *t, P* (*t*), is given by the equation

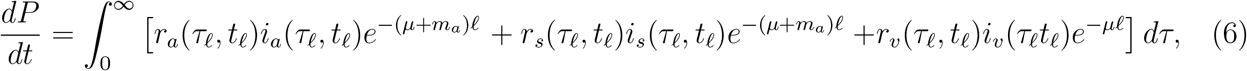

with initial condition *P* (0) = 0.

The cumulative number of SARS-CoV-2 naive individuals who have reached full vaccination status by time *t* is given by

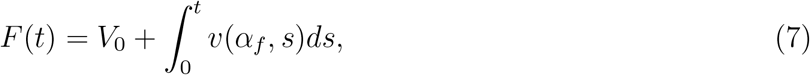

and the cumulative number of previously infected individuals reaching full vaccination status by time *t* is given by

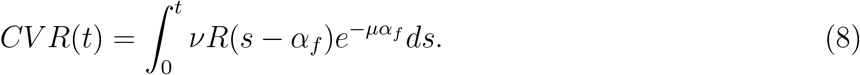

Thus the cumulative number of fully vaccinated individuals at time *t* is

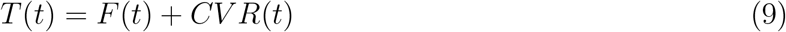

Breakthrough cases are infections of fully vaccinated individuals, so the cumulative number of breakthrough cases up to time *t, B*(*t*), is the number of new infections occurring in individuals with *α* ≥ *α*_*f*_. *B*(*t*) is given by the equation

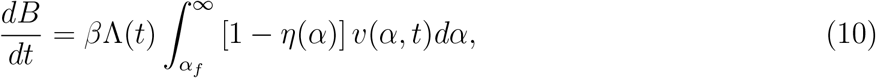

with intitial condition *B*(0) = 0.

### 2.2 Parameter choices

We parametrize our model based on efficacy data from Pfizer-BioNTech vaccine against the alpha and delta variants [4,28]. Therefore, we set the rate 0 ≤ *η*(*α*) ≤ 1 describing the degree of protection against the alpha variant, *α* days after vaccine administration, to

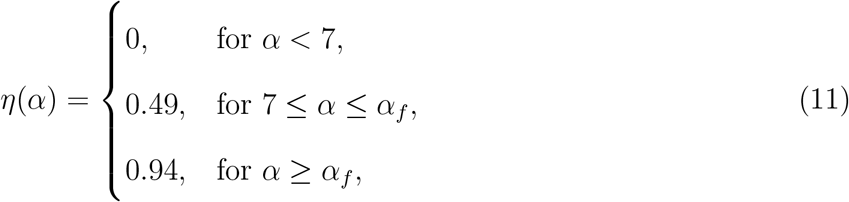

and against delta variant, *α* days after vaccine administration, to

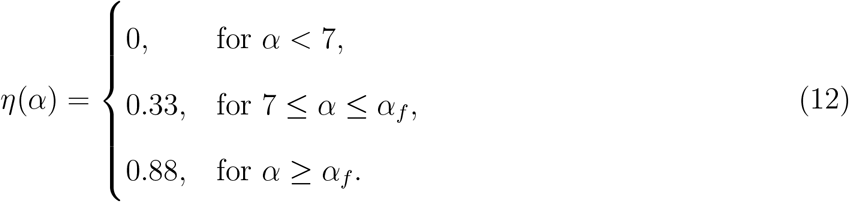

The CDC defines a fully vaccinated individual as one who is ≥ 14 days past receipt of all recommended doses of an FDA-authorized vaccine [12]. In our model, this corresponds to *α* ≥ *α*_*f*_ = 35 days. Levine et al. have shown that, for the first 11 days following the Pfizer-BioNTech vaccination, the cycle threshold (CT) values in infected vaccinated individuals do not change compared to those of infected yet non-vaccinated individuals [27]. We therefore set *α*_*v*_ = 11 days. Afterwards, virus levels decrease by four-fold in vaccinated individuals [27]. The reduction in virus load leads to both shorter infectivity period, ranging between 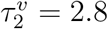 and 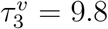 days post infection, and shorter time for detection by tests, ranging between 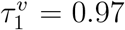 and 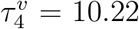 days post infection for RT-PCR (see figure 1). We assume a PCR test return delay of *ℓ* = 1 days.

**Figure 1:**
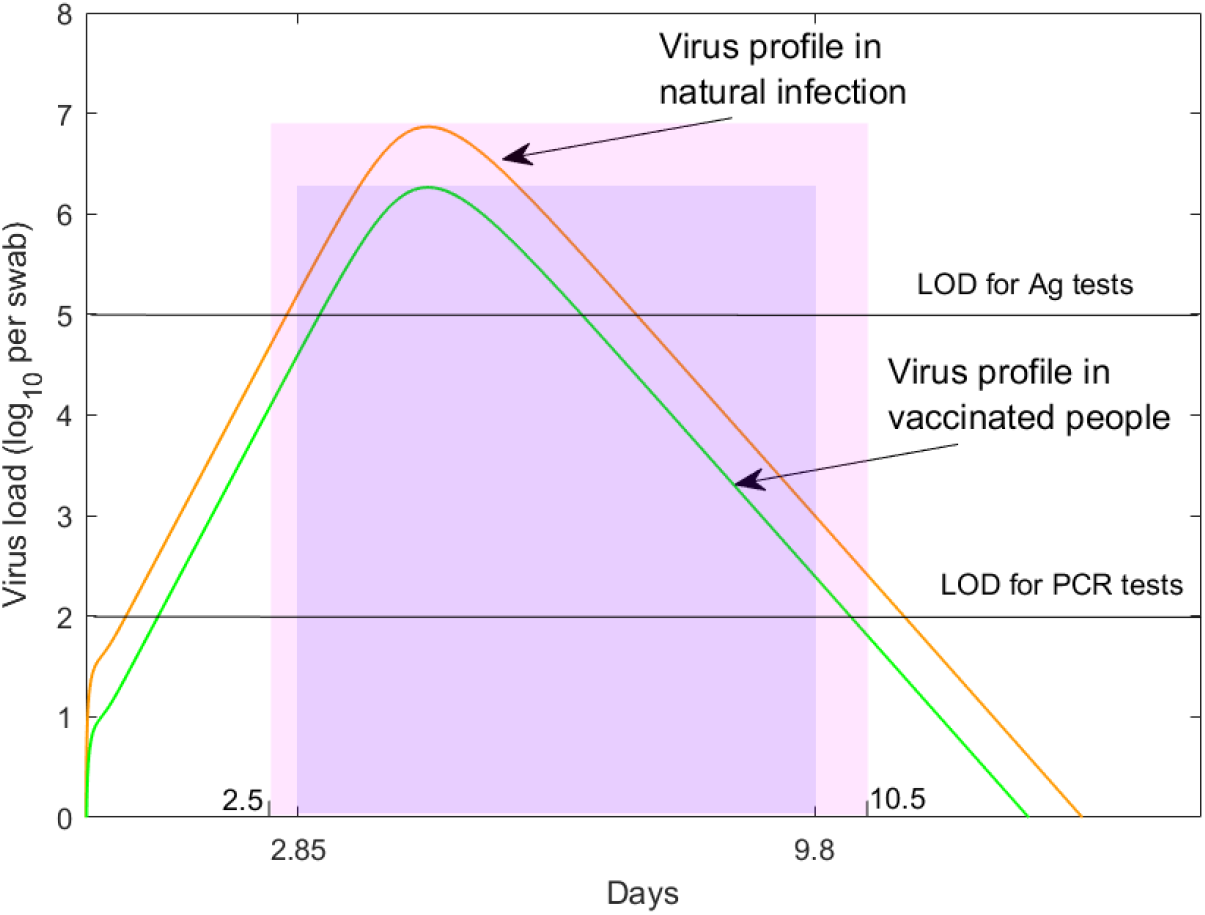
Virus profiles in non-vaccinated and vaccinated individuals. log_10_ virus load per swab over time during alpha variant natural infection (red line) and vaccination (green line) as given by the within-host model in [24]. Non-vaccinated patients are assumed to be infectious from *t* = 2.5 days till *t* = 10.5 days (shaded pink region). Vaccinated patients are assumed to be infectious from *t* = 2.85 days till *t* = 9.8 days (shaded purple region). Black horizontal lines correspond to RT-PCR test detection threshold (LOD) of log_10_(*V*) = 2 per swab and antigen test detection threshold (LOD) of log_10_(*V*) = 5 per swab.

The other parameters *{β, µ, m*_*a*_, *m*_*s*_, *f, τ*_*q*_*}* are as in our previous study [16]. A summary of parameters is given in table 1.

**Table 1:** Parameter values and initial conditions used in model eq. (2). * values account for the delta variant.

| Fixed parameters | Description | Value | Source |
| --- | --- | --- | --- |
| $\beta$ | Transmission rate | 0.25 (0.4*)/day | |
| $b$ | Birth rate | $7 \times 10^{-4}$/day | [34] |
| $\mu$ | Death rate | $7 \times 10^{-4}$/day | [34] |
| $m_j$ | Disease induced mortality rate | $10^{-3}$/day | |
| $f$ | Fraction of symptomatic infections | 0.7 | [21] |
| $\gamma$ | Relative asymp. infectiousness | 0.7 | |
| $\alpha_v$ | Age of vaccination for reduced virus | 11 days | [27] |
| $\alpha_f$ | Age of vaccination corresponding to full protection | 35 days | [5] |
| $\tau_q$ | Recovery age | 14 days | [16] |
| $\ell$ | PCR test return delay | 1 day | |
| $\tau_j^1$ | Age of onset of virus detectability | 0.554 days | |
| $\tau_j^2$ | Age of onset of infectiousness in non-vaccinated | 2.5 days | [41] |
| $\tau_j^2$ | Age of onset of infectiousness in vaccinated | 2.8 days | [27] |
| $\tau_j^3$ | Age of end of infectiousness in non-vaccinated | 10.5 days | [21, 22] |
| $\tau_j^3$ | Age of end of infectiousness in vaccinated | 9.8 days | [27] |
| $\tau_j^4$ | Age of loss of virus detectability | 10.95 days | |
| $\nu$ | Vaccination rate | varied | |
| $t_1$ | Time when additional vaccination is initiated | 20 days | |
| $C$ | testing capacity | varied | |
| Initial conditions | Description | Value | Source |
| $S(0)$ | Susceptible population | $0.95 - V(0)$ | |
| $V(\alpha, 0) = V_0$ | Vaccine prevalence | varied | |
| $i_s(\tau, 0)$ | Infected symptomatic population | $0.05f\delta(\tau)$ | |
| $i_a(\tau, 0)$ | Infected asymptomatic population | $0.05(1 - f)\delta(\tau)$ | |
| $i_v(\tau, 0)$ | Infected vaccinated population | 0 | |
| $R(0)$ | Recovered population | 0 | |
| $R_{v1}(0)$ | Vaccinated after natural infection | 0 | |
| $R_{v2}(0)$ | Vaccinated, infected, and recovered | 0 | |

## 3 Results

### Alpha variant dynamics in the absence of testing

To simulate an initially undetected outbreak in a partially vaccinated population, we assume that at time *t*_0_ = 0, when a total of *V* (0) = *V*_0_ = 30% of individuals have been fully vaccinated, *I*_0_ = 5% of the population is infected with the alpha variant, *f* = 0.7 with symptomatic and 1 − *f* = 0.3 with asymptomatic disease. The vaccine efficacy is given by (11), *v* = 1% additional vaccines are administered daily starting with day *t*_1_ = 20, and no testing is considered. Daily symptomatic, asymptomatic and breakthrough cases peak at 10%, 4.5% and 0.43% of the entire population at days 13, 13 and 46, respectively (see figure 2A, left panel, red, blue and cyan lines). At day 100, a cumulative Σ_*noTests*_(100) = 37.9% of the population has been infected when not fully vaccinated (see figure 2A, middle panel, magenta line) and a cumulative *B*(100) = 1.65% of the population has been infected while fully vaccinated (see figure 2A, middle panel, green line). Lastly, at day 100, a cumulative *F* (100) = 42.6% of the naive population has been fully vaccinated (see figure 2A, right panel, cyan line), a cumulative *CV R*(100) = 9.8% of the population has been fully vaccinated after recovering from natural infection (see figure 2A, right panel, magenta line), and a cumulative *T* (100) = 52.4% of the population has been vaccinated (see figure 2, right panel, black line). A quantity of interest is the percent breakthrough number, *B*(100)*/F* (100), defined as the percent of cumulative naive fully vaccinated individuals that get infected divided by the cumulative fully vaccinated population. In this case, we obtain a percent breakthrough case rate *B*(100)*/F* (100) = 3.88% at day 100.

**Figure 2:**
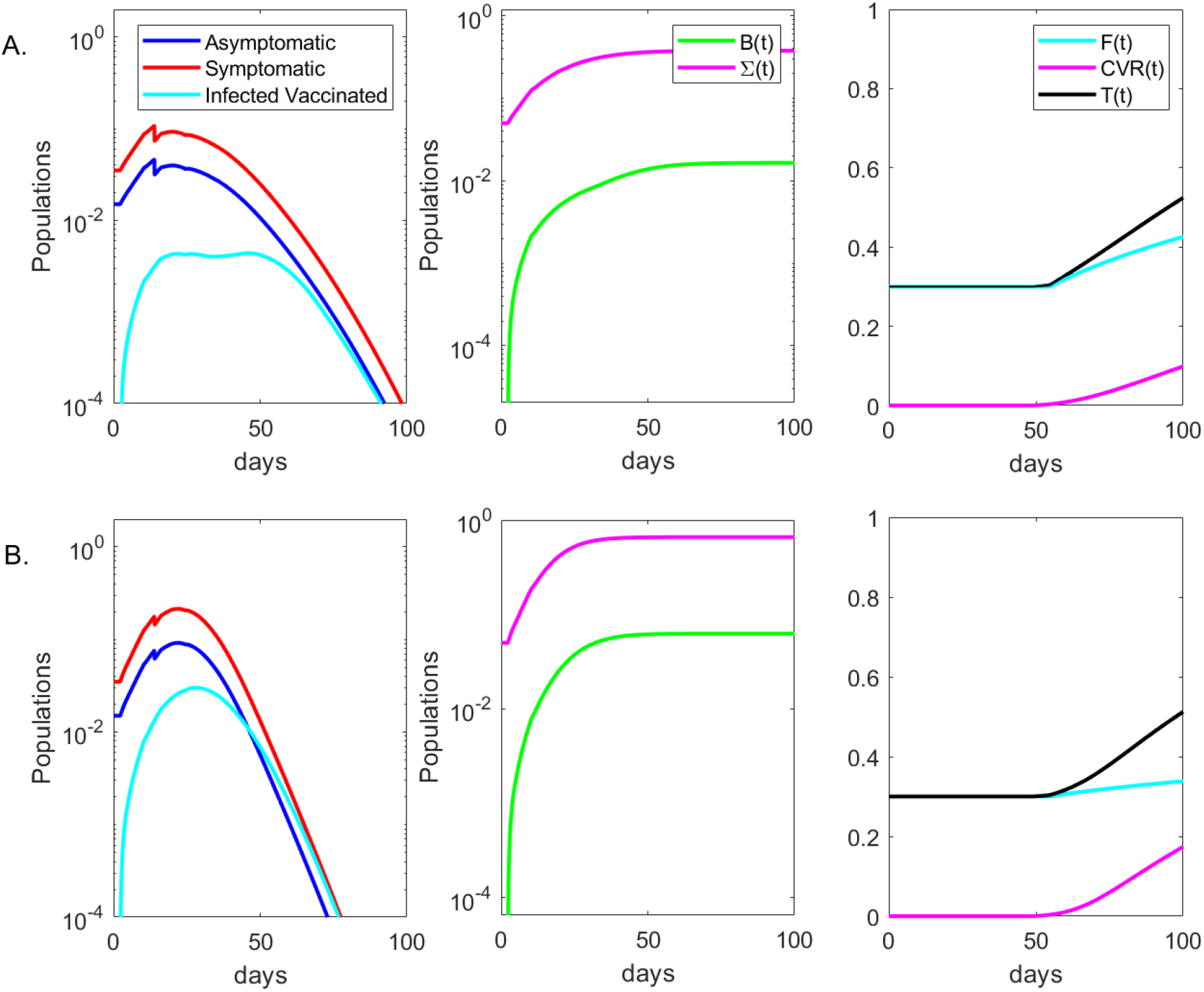
Dynamics of alpha and delta variants infection over time. Left panels: daily asymptomatic (blue), symptomatic (red), and breakthrough (cyan) infections over time; Middle panels: cumulative cases Σ(*t*) (magenta), cumulative breakthrough cases *B*(*t*) (green) over time; Right panels: cumulative fully vaccinated *F* (*t*) (cyan), cumulative vaccinated after infection *CV R*(*t*) (magenta) and cumulative total vaccinated *T* (*t*) (black) over time in the absence of testing. Panel A: alpha variant; Panel B: delta variant. The background vaccination is 30% and the other parameters and initial conditions are given in Table 1.

To more closely determine the relationship between the percent breakthrough cases at day 100, *B*(100)*/F* (100), the vaccination level at the start of the outbreak, *V*_0_, and daily vaccination level starting at day 20, *v*, we derive a heat map for the percent breakthrough cases for smaller *V*_0_ and *v* increments (see Figure 3 A). We observe that the percent breakthrough cases range between 0.7% for *V*_0_ = 80% and *v* = 5% and 6.45% for *V*_0_ = 10% and *v* = 1% (see Figure 3 A). Having a large percent of the population vaccinated at the time of the outbreak results in a decrease in the proportion of fully vaccinated individuals experiencing breakthrough cases.

**Figure 3:**
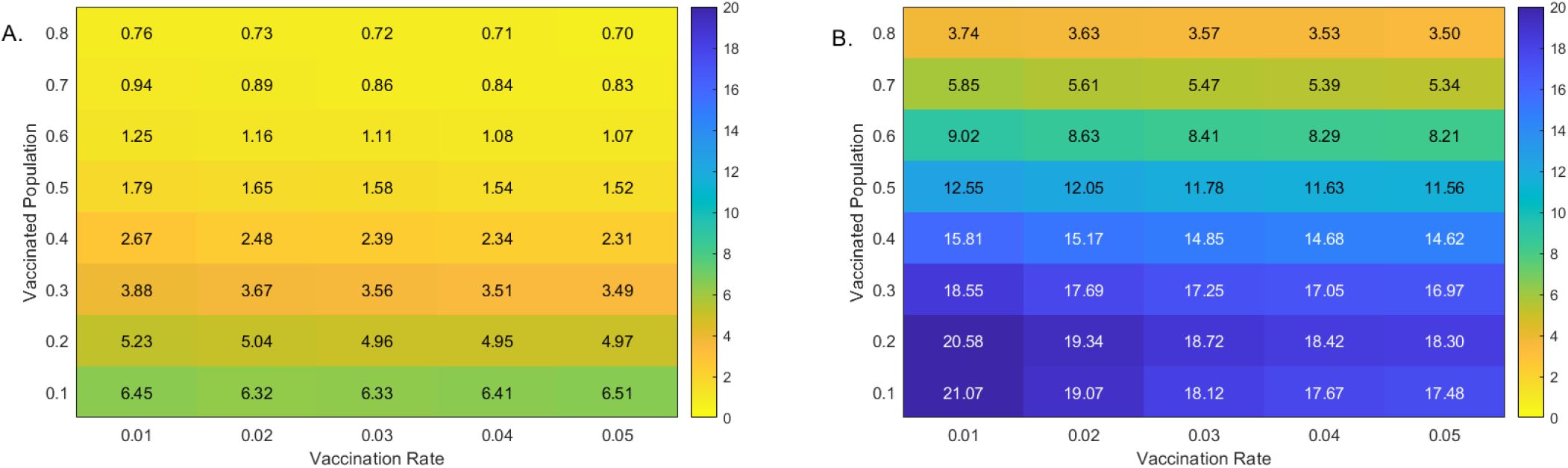
Percent breakthrough cases at day 100. Heatmaps for the percent breakthrough cases in the vaccinated population at day 100 versus additional daily vaccines, *v*, and background vaccination levels, *V*_0_. Panel A: alpha variant; Panel B: delta variant. Parameters and initial conditions are given in Table 1.

### Alpha variant model outcomes in the presence of testing

We investigate the effect of fixed daily testing with capacity *C*, administrated beginning at day *t*_1_ = 20 in two cases. Case 1: only the non-vaccinated group is tested; Case 2: both the vaccinated and non-vaccinated groups are tested. In particular, we quantify the effects of RT-PCR tests with return delay of one day in reducing total infections 100 days after the outbreak, Σ_*noTest*_(100) − Σ_*Test*_(100). For low vaccinated population, increased testing results in increased reduction in cumulative cases, regardless of the testing strategy. In particular, for *V*_0_ = 10% vaccinated population and no testing, the outbreak results in Σ_*noTests*_(100) = 62.8% infections by day 100 (see figure 4, grey heatmaps). When testing only non-vaccinated individuals, this number is reduced by 5.9%, 16.3% and 28.1% for *C* = 0.03, *C* = 0.1 and *C* = 0.5, respectively (see figure 4A, colored heatmaps). When testing everyone, this number is reduced by 5%, 14.7% and 28% for *C* = 0.03, *C* = 0.1 and *C* = 0.5, respectively (see figure 4B, colored heatmaps).

**Figure 4:**
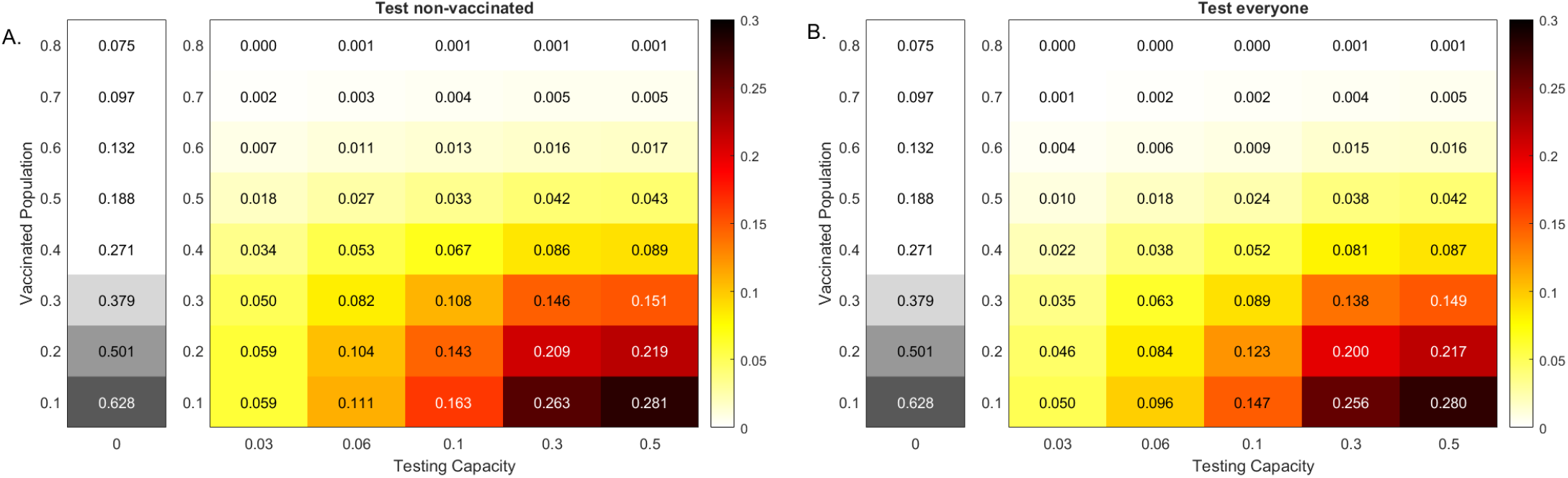
Reduction in alpha variant cases at 100 days. Heatmaps for the reduction in cumulative cases at 100 days after an outbreak with an alpha variant, Σ_*noTests*_(100) − Σ_*Tests*_(100), as given by model eq. (2) versus RT-PCR testing capacity with a return delay of 1 day, *C*, and background vaccination levels, *V*_0_. Panel A: Test non-vaccinated only; Panel B: Test everybody. The gray heatmaps represent the cumulative cases at day 100 in the absence of testing, Σ_*noTest*_(100). Parameters and initial conditions are given in Table 1.

For high vaccinated population, increased testing has no effect on cumulative cases, regardless of the testing strategy. In particular, for *V*_0_ = 80% vaccinated population and no testing, the outbreak results in Σ_*noTests*_(100) = 7.5% infections by day 100 (see figure 4, grey heatmaps). Testing affects this number by 0.1% for *C* ≥ 0.06 when testing non-vaccinated individuals and for *C* ≥ 0.3 when testing everyone, respectively (see figure 4, colored heatmaps).

Note that, for both testing scenarios, the amount of reduction decreases with the increase of *V*_0_ (for a fixed *v*) and increases with the increase of testing capacity *C* (for a fixed *V*_0_). Moreover, the highest decrease happens for the highest testing in the least vaccinated population. Lastly, in all cases, we predict that testing only non-vaccinated results in improved outcomes.

### Delta variant dynamics in the absence of testing

We examine the effect of the Pfizer–BioNTech vaccine on blocking infections with the delta variant. To account for the increased infectiousness of delta, we increase the transmissibility rate to *β* = 0.4, 1.5-times higher than in the case of the alpha variant. Moreover, we modify the Pfizer–BioNTech effectiveness to (12).

As before, we simulate an initially undetected outbreak in a partially vaccinated population, with *V*_0_ = 30% of the population fully vaccinated at the time of the outbreak, *v* = 1% additional vaccines are administered daily starting with day *t*_1_ = 20, and no testing is considered. Daily symptomatic, asymptomatic and breakthrough cases peak at 21.4%, 9.1% and 3% of the entire population at days 22, 22 and 28.5, respectively (see figure 2B, left panel, red, blue and cyan lines). At day 100, a cumulative Σ_*noTests*_(100) = 66% of the population has been infected when not fully vaccinated (see figure 2B, middle panel, magenta line) and a cumulative *B*(100) = 6.27% of the population has been infected while fully vaccinated (see figure 2B, middle panel, green line). Lastly, at day 100, we predict that a cumulative *F* (100) = 33.8% of the naive population has been fully vaccinated (see figure 2B, right panel, cyan line), a cumulative *CV R*(100) = 17.3% of the population has been fully vaccinated after recovering from natural infection (see figure 2B, right panel, magenta line), and a cumulative *T* (100) = 51.2% of the population has been vaccinated (see figure 2B, right panel, black line). The percent breakthrough case rate at day 100 becomes *B*(100)*/F* (100) = 18.55%.

When we expand our analysis to include smaller *V*_0_ and *v* increments, we predict that the breakthrough cases at day 100 increase to *B*(100)*/F* (100) = 21.07% (compared to 6.45% for the alpha variant) for *V*_0_ = 10% and *v* = 1% and to *B*(100)*/F* (100) = 3.5% (compared to 0.7% for the alpha variant) for *V*_0_ = 80% and *v* = 5% (see Figure 3 B). For this more contagious variant, having a large percent of the population vaccinated at the time of the outbreak significantly decreases the breakthrough cases.

### Delta variant model outcomes in the presence of testing

We next investigate the effect of fixed daily testing with capacity *C*, administered at day *t*_1_ = 20 in reducing total infections 100 days after the outbreak, Σ_*noTest*_(100) − Σ_*Test*_(100), when only the non-vaccinated and when everyone is tested.

When less than 60% of the population has been vaccinated at the time of the outbreak, increased testing leads to increased reduction in cumulative cases, regardless of the testing strategy. In particular, for *V*_0_ = 10% vaccinated population and no testing, the outbreak results in Σ_*noTest*_(100) = 85.4% of the population being infected by day 100 (see figure 5, grey heatmaps). When testing non-vaccinated individuals only, this number is reduced by 1.9%, 6.6% and 18.6% for *C* = 0.03, *C* = 0.1 and *C* = 0.5, respectively (see figure 5A, colored heatmaps). When testing everyone, this number is reduced by 1.7%, 5.9% and 18.5% for *C* = 0.03, *C* = 0.1 and *C* = 0.5, respectively (see figure 5B, colored heatmaps). For *V*_0_ = 30% and no testing, the outbreak results in Σ_*noTest*_(100) = 66% of the population being infected by day 100 (see figure 5, grey heatmaps). When testing non-vaccinated individuals only, this number is reduced by 3.6%, 10.6% and 20% for *C* = 0.03, *C* = 0.1 and *C* = 0.5, respectively (see figure 5A, colored heatmaps). When testing everyone, this number is reduced by 2.6%, 8.2% and 20% for *C* = 0.03, *C* = 0.1 and *C* = 0.5, respectively (see figure 5B, colored heatmaps). Finally, for *V*_0_ = 60% vaccinated population and no testing, the outbreak results in Σ_*noTest*_(100) = 30.4% of the population being infected by day 100 (see figure 5, grey heatmaps). When testing only non-vaccinated individuals, this number is reduced by 3.9%, 7.4% and 9% for *C* = 0.03, *C* = 0.1 and *C* = 0.5, respectively (see figure 5A, colored heatmaps). When testing everyone, this number is reduced by 2.1%, 5.2% and 9.2% for *C* = 0.03, *C* = 0.1 and *C* = 0.5, respectively (see figure 5B, colored heatmaps). As with the alpha variant, the amount of reduction increases with the increase of *C* (for a fixed *V*_0_), regardless of the testing scenarios. Unlike the alpha variant case, however, the reduction does not decrease monotonically with the increase of *V*_0_ (for fixed *C*). The maximum reduction occurs at different *V*_0_ values for different testing capacities *C*. In particular, for both testing strategies, the maximum reductions occur at *V*_0_ = 40% for *C* = 0.1 and *V*_0_ = 30% for *C* = 0.3.

**Figure 5:**
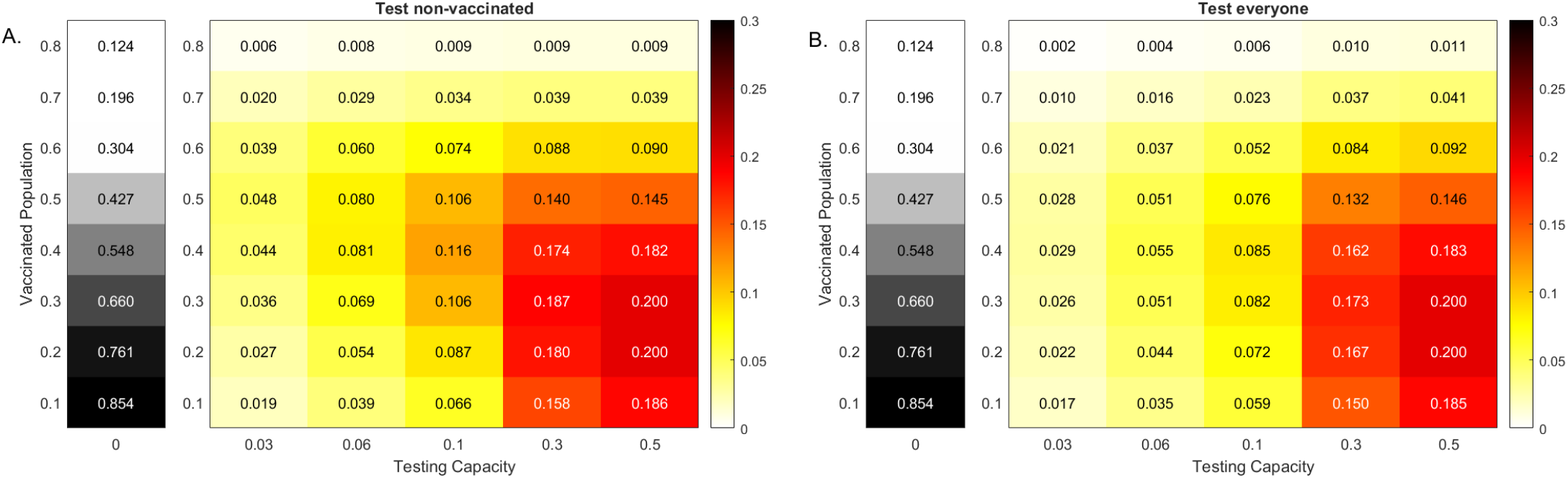
Reduction in delta variant cases at 100 days. Heatmaps for the reduction in cumulative cases at 100 days after an outbreak with a delta variant, Σ_*noTests*_(100) − Σ_*Tests*_(100), as given by model eq. (2) versus RT-PCR testing capacity with a return delay of 1 day, *C*, and background vaccination levels, *V*_0_. Panel A: Test non-vaccinated only; Panel B: Test everybody. The gray heatmaps represent the cumulative cases at day 100 in the absence of testing, Σ_*noTest*_(100). Parameters and initial conditions are given in Table 1.

When more than 60% of the population is vaccinated, increased testing results in increased reduction in cumulative cases when we test everyone, and stops being effective when we only test non-vaccinated. In particular, for *V*_0_ = 80% vaccinated population, testing only vaccinated for *C* ≥ 0.1 does not improve the outcomes (see figure 5, colored heatmaps). That is why, for high vaccinated populations, testing everyone leads to a slight increase in case reductions.

## 4 Discussion

Reaching herd-immunity to SARS-CoV-2 through vaccination and natural infection is made harder by vaccine hesitancy, vaccine shortage around the world, emergence of new variants, and delayed vaccination approval for children. Therefore, additional public health interventions such as masks, social distance, and surveillance testing are still needed. In previous work, we have used mathematical models to show that surveillance testing can be an effective public health intervention to reduce a SARS-CoV-2 outbreak in a non-vaccinated population [16]. The significance of surveillance testing needs to be re-evaluated in the context of vaccine prevalence, emergence of new variants, and waning immunity. CDC recommends that vaccinated people get tested only when experiencing COVID-19 symptoms or came into close contact with someone with COVID-19 [2]. To better determine the role of surveillance tests in a rapidly changing environment, we developed a multi-scale mathematical model of COVID-19 transmission in a mixed vaccinated and non-vaccinated population. Our model investigates how the prevalence of vaccination impacts the effectiveness of testing. We compare two testing strategies: one in which tests are administered to both vaccinated and non-vaccinated individuals and one in which tests are administered to non-vaccinated individuals alone. Additionally, we separately consider both the alpha and delta variants of the virus.

In the case of the alpha variant, where vaccines are highly effective in blocking transmission, we find that testing remains an effective intervention when the overall prevalence of vaccination is low to moderate. For higher vaccination levels, the impact of testing is diminished, even relative to the smaller outbreaks that occur in those scenarios. For any fixed testing capacity, the number of cases prevented decreases with increasing vaccine prevalence.

For the delta variant, where vaccine efficacy in blocking transmission is reduced, a more complex pattern of testing effectiveness is apparent. For low vaccination, the impact of testing is low, as testing is not sufficient to overcome the force of infection created by the delta variant. This difference to the case of the alpha variant is a result of the magnitude of the outbreak, which is driven by increased transmissibility and the reduced effectiveness of the vaccine against the delta variant. Interestingly, as the prevalence of vaccination increases, the number of cases prevented by testing increases as well, even though the number of cases that would occur in the absence of testing declines. As vaccination prevalence increases further, the effectiveness of a fixed testing capacity declines again, due to the significant reduction in the number of cases expected in the absence of testing, as in the case of the alpha variant. Thus, when considering the delta variant, the impact of a fixed testing capacity is highest for moderate vaccination prevalence and lower for low and high vaccination prevalence.

We investigated differences in testing strategies. In the United States, it is common for surveillance testing to focus on the non-vaccinated population [2]. Here we compare the differences in outcomes for testing strategies that include only the non-vaccinated and those that include both vaccinated and non-vaccinated populations. We find that testing strategies that focus on the non-vaccinated population are generally more effective than broad testing strategies. For the alpha variant, this is true for all cases considered. For the delta variant, a broad testing strategy is preferable when the testing capacity exceeds the non-vaccinated population, a result that has been reported in other modeling studies [32]. This indicates that the most effective strategy should focus first on testing the non-vaccinated population, then use excess capacity in the non-vaccinated population.

As expected, for fixed vaccination prevalence, increasing testing capacity increases the number of cases prevented. However, for high vaccination prevalence, the impact of additional testing eventually saturates, and further testing has little or no effect.

We have also studied the prevalence of infection within the vaccinated population, so-called “breakthrough” cases. For the alpha variant, the prevalence of breakthrough cases is uniformly low (ranging between 0.7% and 6.45%), and decreases as the population vaccination prevalence increases. These results are similar to those from clinical studies which reported an alpha variant incidence of breakthrough infections of 0.5% in health care workers in US who received either the Pfizer–BioNTech or the Moderna vaccine [19], of 2.6% in health care workers in Israel who received both doses of Pfizer–BioNTech vaccine [8], and of 0.08% in New York metropolitan area vaccinated with either Pfizer–BioNTech, Moderna, or Janssen vaccines [14]. For the delta variant, our model predicts that breakthrough cases are much more prevalent in the vaccinated population and range between 3.5% and 21.07%. These are comparable with the 8.4% breakthrough cases reported in Houston hospitals [33] and lower than the 28% reported in the DC area [29]. While the increased transmissibility of the delta variant and the decreased effectiveness of vaccine are necessary conditions for this increase in breakthrough cases, the primary driving factor is the extent of the outbreak in the non-vaccinated population. As the vaccinated population becomes larger, both the total number of breakthrough cases and their prevalence decrease.

It is important to note that, in this study, the measure for the effectiveness of testing is the prevention of transmission, not the prevention of disease, hospitalization, or death. The true transmission is generally not known in an ongoing outbreak, even when testing is widespread. Public health outcomes and vaccine effectiveness are generally expressed in terms of preventing disease, with all three vaccines approved for use in the United States being highly effective in preventing hospitalization and deaths regardless of the variant [31]. While preventing disease is the immediate goal, we argue that preventing transmission even when a large fraction of the population has been vaccinated (through surveillance testing) is a worthy long-term goal that may allow emergent variant strains resistant to vaccination to go extinct before becoming the next dominant strain [37]. We have started the outbreak with a large number of infectious individuals. If we decrease that initial burden from 5% to 1%, the epidemic curve flattens and the time to the peak lengthens. As a result, the effect of testing in lower vaccinated populations increases significantly (not shown). As before, the effect of testing in the highly vaccinated population saturates. This indicates that early detection of nascent outbreaks is essential for the effectiveness of testing as a public health intervention.

Our study is limited to PCR testing with a return delay of one day. In the past, we have investigated the trade-off between employing faster, cheaper, yet less sensitive antigen tests at a more frequent rate. When the same testing capacity is used, the antigen tests underperform the PCR tests with one day delay (not shown). This is due to decreased interval of detection for the antigen tests (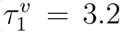 and 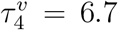 days post infection). If the frequency of antigen test administration is increased, or in places where the PCR return is long, antigen test can present a reliable alternative for surveillance testing.

## 5 Conclusions

In summary, we have developed a multi-scale model of SARS-CoV-2 transmission in a vaccinated population. We found that, when the alpha variant is dominant, testing is effective when vaccination levels are low to moderate and its impact is diminished when vaccination levels are high. When the delta variant is dominant, widespread vaccination is necessary in order to prevent significant outbreaks. When only moderate vaccination prevalence can be achieved, frequent testing can significantly reduce the cumulative size of the outbreak, and the impact of testing is greatest when it is focused on the remaining non-vaccinated population.

## Data Availability

All data produced in this study are contained in the manuscript.

